# Detection and quantification of hepatitis A virus titers from wastewater in South Africa and comparison with clinical data from the National Surveillance Database

**DOI:** 10.1101/2024.12.19.24318086

**Authors:** Kathleen Subramoney, Sipho Gwala, Emmanuel Phalane, Mokgaetji Macheke, Natasha Singh, Thabo Mangena, Nkosenhle Ndlovu, Nosihle Msomi, Sibonginkosi Maposa, Chenoa Sankar, Fiona Els, Phindile Ntuli, Mantshali Motloung, Victor Mabasa, Kerrigan McCarthy, Mukhlid Yousif

## Abstract

Wastewater surveillance is useful for monitoring the prevalence of hepatitis A virus (HAV). We developed and optimized HAV detection and quantification methods for wastewater samples, and applied them to samples collected through a national wastewater surveillance program. Previously identified 5’-untranslated region-targeting primers and probes were used to develop the assay. Serial dilutions of HAV-positive clinical samples were used to validate and determine limits of quantification (LOQ). Retrospective testing of weekly wastewater samples collected through the SARS-CoV-2 wastewater surveillance program at 26 sites in Gauteng (August 2021 to March 2024) were undertaken using ultrafiltration-based concentration, and nucleic acids were extracted using the KingFisher Flex purification system with a wastewater isolation kit. A digital PCR assay was used for HAV detection and quantification (as genome copies/μL). Clinical data from the Surveillance Database Warehouse of the National Health Laboratory Service were compared with wastewater data, epidemiological week-wise and district-wise, to determine correlations between the datasets. Based on the validation results, one partition on the digital PCR (dPCR) platform was equivalent to an LOQ of 0.4 genome copies/μL. In total, 2013 wastewater samples were tested, of which 349 were positive for HAV (17.3%), wherein the majority (304, 87.1%) had the lowest HAV concentration (2.0-2.7 gc/μL, 1-5 partitions), followed by 20 samples (5.7%) with concentrations of 2.8-3.0 gc/μL (6-10 partitions). HAV was detected in 17.1% (241/1170) of the Gauteng samples, and a 26.1% correlation between anti-HAV IgM in clinical samples and HAV in wastewater samples was detected. We successfully developed and optimized a dPCR method to detect and quantify HAV in wastewater samples, and determined its LOQ. Further analysis is required to compare the wastewater data with clinical surveillance data to facilitate appropriate interpretation of the results.

## 1. Introduction

Hepatitis A virus (HAV) is primarily transmitted via the fecal-oral route, and causes mild to severe symptoms, including vomiting, diarrhea, fever, dark-colored urine, and jaundice, as well as fulminant hepatitis [1–3]. Children under 5 years of age either present with mild symptoms or are asymptomatic whereas adults present with mild to severe symptoms [1–3]. HAV incidence is usually higher in low-income countries because of poor sanitation, contaminated water, and poor sewage disposal, with consequent contamination of food or water [4–6]. Recent studies indicate a decline in HAV incidence in middle-to high-income countries, and outbreaks are commonly associated with high-risk groups, including travelers from highly endemic regions, men who have sex with men (MSM), the administration of injectable drugs, and homeless people with poor sanitary habits [4–6]. In countries with high HAV endemicity, infection occurs during early childhood, and immunity develops with positivity for anti-HAV immunoglobulin (IgG) by adulthood. Unlike in countries with low HAV endemicity, adults are susceptible to HAV and are often symptomatic [7]. However, because HAV-infected children are usually asymptomatic and only seek medical treatment and testing upon the presentation of symptoms, the prevalence of the disease may be underestimated.

Low-income regions suffer from the most severe threat of hepatitis A, with the highest age-standardized incidence rates of HAV in sub-Saharan Africa [4]. In South Africa, where HAV disease contributes to approximately 0.3% of deaths annually, and the severity of the disease increases with age [8], HAV infection is considered a notifiable medical condition (NMC) in the surveillance system, based on passive surveillance. In this approach, reports on cases are notified by clinicians and laboratories that test for the virus under the National Health Laboratory Services (NHLS), and are used to determine the thresholds for public health action [9]. However, the data from the private sector currently remain unreported. The environmental presence of the HAV in low- and middle-income countries (LMICs) has been closely studied [10]. Active surveillance of HAV in South Africa is imperative for the routine monitoring of cluster influx through the implementation of public health interventions to prevent outbreaks [9]. Studies in the Western Cape and Gauteng provinces reported <90% HAV seropositivity in children aged 10 years, which indicates a transition from high to intermediate endemicity in South Africa [11,12]. Although the World Health Organization (WHO) recommends that HAV vaccination be integrated into national immunization schedules for children in African populations where seroprevalence drops to <90% by age 10, this approach it is not considered cost-effective for routine use in the public health sector [13].

Wastewater surveillance has been implemented globally for the detection of viruses, including severe acute respiratory syndrome coronavirus 2 (SARS-CoV-2) and poliovirus, in conjunction with routine clinical surveillance for in-depth screening in the population of interest [14, 15]. This is an effective early warning system for the detection of emerging and circulating viruses, especially with the knowledge that younger children are asymptomatic and will not seek healthcare services [16–18] As HAV is stable under diverse environmental conditions, including high temperatures (up to 80°C) and a low pH range (1-2), its presence is detectable in fresh and wastewater [2,19, 20]. In South Africa, HAV nucleic acids have been detected in freshwater from selected rivers, dams, and wastewater [21, 22]. Wastewater containing human excretions can enable public health authorities to identify HAV outbreak hotspots, monitor currently circulating strains, study trends in several regions, and facilitate early detection for more informed public health decision-making [23]. In particular, in low- to middle-income countries with restricted access to appropriate resources, wastewater surveillance can be used to monitor HAV incidence in asymptomatic and symptomatic populations and trace the potential sources of outbreaks. However, the detection of HAV in effluent samples was negligible over the sampling period [22] and remained undetectable in the final effluent sampled immediately after chlorination [24].

Currently, there is no active HAV surveillance in South Africa, and data are available from passive surveillance, wherein only symptomatic cases are recorded. Therefore, HAV wastewater surveillance could provide added value to strengthen surveillance systems to monitor trends in HAV incidence, identify outbreaks, and implement appropriate public health responses and prevention measures. Similarly as for other picornaviruses, the 5’UTR is commonly used for the detection of HAV because of its highly conserved nature, making it less prone to nucleotide changes [25–27]. The 5’UTR is approximately 732 nucleotide bases in length and comprises three pyrimidine-rich (80–90%) regions from positions 99–138, 204– 250, and 711–725 [27]. This constitutes a useful tool for detecting HAV in outbreak settings [28].

South Africa is an upper-middle-income country with a population of 66 million individuals within nine provinces that comprise 52 health districts. Approximately 80% of individuals receive healthcare services, including laboratory diagnostic testing, from public sector clinics, hospital facilities, and the National Health Laboratory Service (NHLS). The National Institute for Communicable Diseases (NICD), a division of the NHLS, is responsible for communicable disease surveillance through various programs, including the legally mandated Notifiable Medical Condition Surveillance System (NMCSS). HAV seroprevalence has been previously described [28]. Wastewater and environmental surveillance (WES) has been conducted by the NICD since 2021, and commenced at national sentinel sites (two large wastewater treatment plants in each province). However, limited data on the use of WES for HAV monitoring have been reported in South Africa.

We aimed to optimize methods for HAV detection and quantification from wastewater samples based on the 5’ UTR region. We adopted a digital PCR method that offers sensitive and absolute quantification without the need for a reference and has a greater tolerance to inhibitors present in wastewater samples [29]. We applied the novel method to analyze the samples collected as part of a national wastewater surveillance program, from three “case-study” areas in Gauteng Province, each with 6 sampling points located within the catchment area of a single large wastewater treatment site. Moreover, we compared the findings of the WES data with clinical HAV surveillance data from health facilities located in the same regions.

## 2. Methods

### 2.1. Optimization using real-time reverse-transcription PCR (RT-PCR) detection of HAV

RNA was extracted using the QIAamp RNA Mini Extraction Kit (Qiagen, Hilden, Germany) following the spin column method, according to the manufacturer’s instructions. A one-step real-time RT-PCR assay was used to optimize the primers (HAV68F TCACCGCCGTTTGCCTAG and HAV240R GGAGAGCCCTGGAAGAAAG) and probe (HAV150Probe TTAATTCCTGCAGGTTCAGG labelled with FAM-MGB) for the detection of 5’UTR HAV [30]. The QuantiFast® Pathogen RT-PCR assay (Qiagen, Hilden, Germany) was optimized by preparing the following reaction mix: 5 μL 5× QuantiFast Pathogen Master Mix, 0.25 μL 100× QuantiFast Pathogen RT Mix, equal volumes of 10 µM primers and probe were pooled together and 2.5 µL of the pool was used for the reaction mix, and with 2.5 μL 10× internal control assay and internal control RNA and 8 μL RNA, made up to a volume of 25 μL with nuclease-free water. Previously sequenced HAV-positive clinical specimens (IgM-seropositive and successfully sequenced by Sanger sequencing) were used to optimize the assay, which was performed on an ABI 7500 real-time PCR platform (Applied Biosystems). The PCR cycling conditions were run according to the manufacturer’s instructions, with a modification of the annealing and extension conditions (60°C for 1 min, for 45 cycles).

### 2.2. Optimization using digital PCR (dPCR) from clinical specimens

The QIAcuity OneStep Advanced Probe RT-dPCR assay (Catalog no. 250132, Qiagen, Hilden, Germany) was performed using the QIAcuity Digital PCR system (Qiagen, Hilden, Germany) according to the manufacturer’s instructions, with the primers and probes used for real-time PCR optimization and the previously extracted RNA. The assay was optimized by preparing the following reaction mix: 3 μL 4× OneStep Advanced Probe Master Mix, 0.12 μL 100× OneStep Advanced RT Mix (Reverse Transcription), equal volumes of 20 µM primers and probe pooled together, of which 0.6 µl was added to the reaction mix, 1.5 μL enhancer GC, 5 μL RNA, and nuclease-free water to make up a total volume of 12 μL. The reaction mix was added to an 8.5k (96 well) nanoplate and loaded into the digital PCR instrument. The PCR cycling conditions were set as per the manufacturer’s instructions, with the following modifications: annealing and extension at 60°C for 1 min, for 45 cycles. The master mix was evenly distributed through 8,500 partitions in each well. Nucleic acids in each partition were quantified by imaging each well after the PCR. Results were interpreted as copies/µL. Previously sequenced positive HAV clinical specimens (IgM seropositive and successfully sequenced) were used as positive controls.

### 2.3. Limit of detection

In accordance with the method described above, the limit of quantification (LOQ) of the optimized assay was determined using clinical specimens that were previously confirmed as IgM seropositive and were successfully sequenced. Two-fold dilutions were prepared from 1:2 to 1:1024, with 8 µL RNA extracted from clinical samples used as the template. The cycling conditions were identical to those described above.

### 2.4. Study site, sample collection, and processing of wastewater samples

Next, 1 L grab samples of untreated raw wastewater were collected bi-weekly, weekly, or monthly from sentinel and “case-study” sampling sites. Figure 1 shows the distribution of the sites from which the wastewater samples were collected. The population in the study sites ranged from 1200 to 950000 people. The samples included a combination of influent from the wastewater treatment works (just after the screens) and street line manholes. Samples from the catchment sites are collected between 9am and 11am. From all other sites samples were collected anytime in the morning by trained plant operator. Samples were transported on ice at 4°C to the NICD. Within 24 hours of receipt, the samples were concentrated by ultrafiltration using Centricon® Plus-70 centrifugal filter units (MERCK, Germany), per the manufacturer’s instructions (31). The concentrate was tested for SARS-CoV-2 in real time as previously described (31). The residual concentrates were stored at −20°C. All experiments were performed by the same person, using the same instrument, materials etc.

**Fig. 1.**
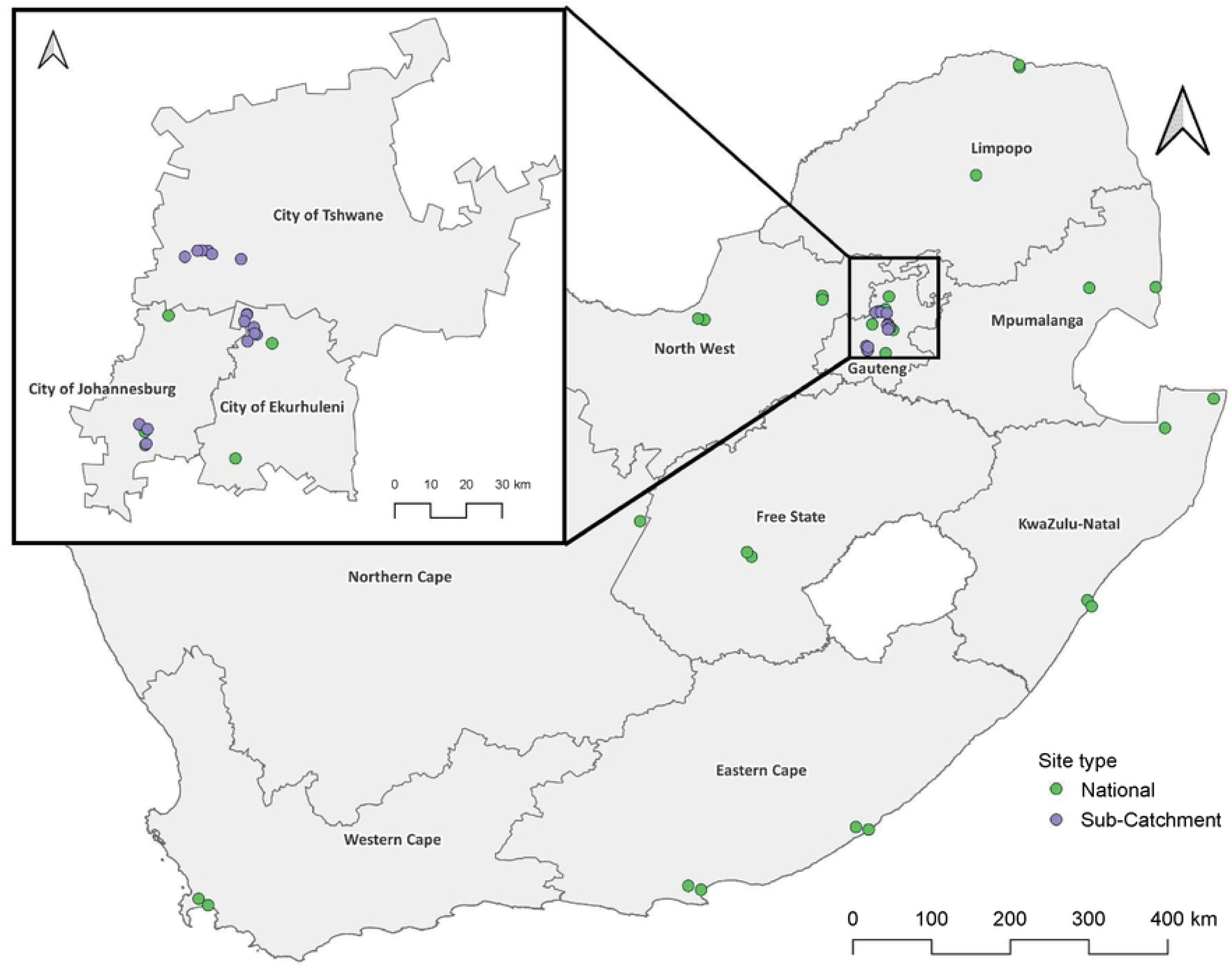
Map of South Africa displaying the wastewater sampling sites in each province. The green circles represent 29 national sampling sites and the purple circles represent 18 community sites within the Gauteng Province.

### 2.5. Total nucleic acid extraction from wastewater samples

Nucleic acids were extracted from the residual concentrate using a MagMAX^TM^ Wastewater Ultra Nucleic Acid Isolation Kit (ThermoFisher Scientific, Massachusetts, United States) on a KingFisher^TM^ Flex instrument (ThermoFisher Scientific, Massachusetts, United States) according to the manufacturer’s instructions.

### 2.6. HAV detection in retained wastewater samples

A duplex assay targeting hepatitis A was performed on all wastewater samples. For each reaction, the mastermix comprised 3 µL 4× OneStep Advanced Probe Master Mix, 0.12 μL 100× OneStep Advanced RT Mix (Reverse Transcription), 0.6 µL 20 µM HAV primer/probe-ROX, 1 µL enhancer GC, with the addition of 8 μL total nucleic acid. The mastermix was briefly centrifuged and 13.82 µL was transferred to its respective well in the 8.5k 96-well dPCR nanoplate, and loaded into the dPCR instrument. The cycling conditions were identical to those described above. The concentration output by the instrument, presented in genome copies/µL, was converted to genome copies/mL using the following formula:

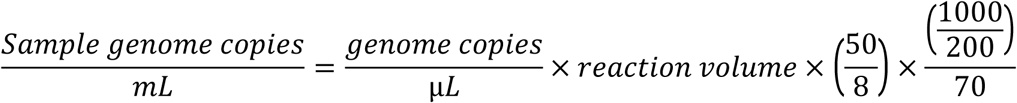

The experiment included a negative control in which no target sample was detected.

### 2.7. Epidemiological analysis of wastewater and clinical HAV surveillance data

As part of the NMC surveillance, the NICD analyzes and interprets laboratory results from routine clinical hepatitis A serology testing conducted at the NHLS across South Africa, as well as from private-sector laboratories. We used anonymized HAV NMC testing data from July 2021 to November 30, 2023, and identified samples from health facilities located in districts where wastewater surveillance testing was conducted. We grouped positive and negative clinical results by epidemiological week and district of the health facility and tallied the positive and negative results for each time-space combination. Similarly, we arranged HAV-positive and HAV-negative wastewater samples according to the epidemiological week and district of the sample collection site. We tallied the positive and negative results for each time–space combination.

We merged the two datasets by epidemiological week and district and compared the wastewater with clinical results. We classified wastewater: clinical pairs as “concordant-positive” when at least one wastewater sample and at least one clinical sample tested positive for HAV in that epidemiological week and in one district. Conversely, if no wastewater samples and no clinical specimens tested positive for HAV in an epidemiological week and district combination, we classified that week-district as ‘concordant-negative.’ The discordant classification was assigned when, in specific week-district combinations, no clinical specimen tested positive but HAV was detected in one or more wastewater samples (discordant-wwpos) or when HAV was detected in at least one clinical specimen but all wastewater samples tested negative for HAV (discordant-wwneg). Furthermore, to account for the extended shedding duration of those infected with HAV both before and after the epidemiological week of diagnosis (assumed to be the date of collection of the clinical specimen), clinical testing from up to 2 weeks before and after the week in which a wastewater sample was tested was considered.

Our rationale for this analytic approach is based on: 1) The epidemiology of HAV infection in a high-endemicity setting, which suggests that when a single positive HAV case is identified through passive surveillance, in all likelihood, there are other asymptomatic, incubating, or undetected cases present in the district; and 2) On account of the inherent and relative insensitivity of wastewater detection for viral RNA targets, a positive HAV result in a wastewater sample indicates the presence of that pathogen in the catchment population, but a negative result cannot confirm the absence of the pathogen; and 3) the need for epidemiologically meaningful analytic approach, from which public health interventions could potentially emerge; and 4) a district as a meaningful unit of analysis for public health interventions.

### 2.8. Statistical analysis

Wastewater testing data were entered directly into RedCap [30, 31], and the data required for hepatitis A were exported and analyzed using Excel. All graphs were generated using Microsoft Excel. Clinical data from the Surveillance Data Warehouse (SDW) were analyzed using the R programming language (insert version of R used). ArcGIS was used to generate a map of South Africa indicating the proportion of wastewater samples that tested positive for HAV and the proportion of anti-HAV IgM from the clinical data. The anti-HAV IgM data from SDW were compared with wastewater data by analyzing the data of the corresponding epidemiological week and district (referred to as epidemiological week-district pairs) based on where the wastewater testing was conducted. A positive epidemiological week–district pair was defined as the presence of at least one wastewater or clinical sample that tested positive for HAV.

### 2.9. Ethics

The NICD conducts all routine clinical surveillance, including NMC surveillance, in a protocol reviewed and approved by the University of the Witwatersrand Human Research Ethics Committee (HREC) M210752 and under the legal authority of the National Health Act (no. 61 of 2003). Wastewater sampling and testing were performed using an HREC system (M230828). Informed consent was not required as we only used anonymized data.

## 3. Results

### 3.1. Development of HAV detection and quantification assays for wastewater samples

Real-time PCR and dPCR for the detection of the HAV 5’ UTR was successfully optimized (Fig. 2). The number of partitions and concentration of genome copies (gc/mL) were compared to determine the LOQ, which was verified as one partition, with a Ct-value of 38.5, and a concentration of 0.4 gc/µL (Fig. 3). When comparing the Ct value to the concentration and the Ct value to the number of partitions (Fig. 3), the R^2^ value was 0.967, which indicated 96% reliability.

**Fig. 2.**
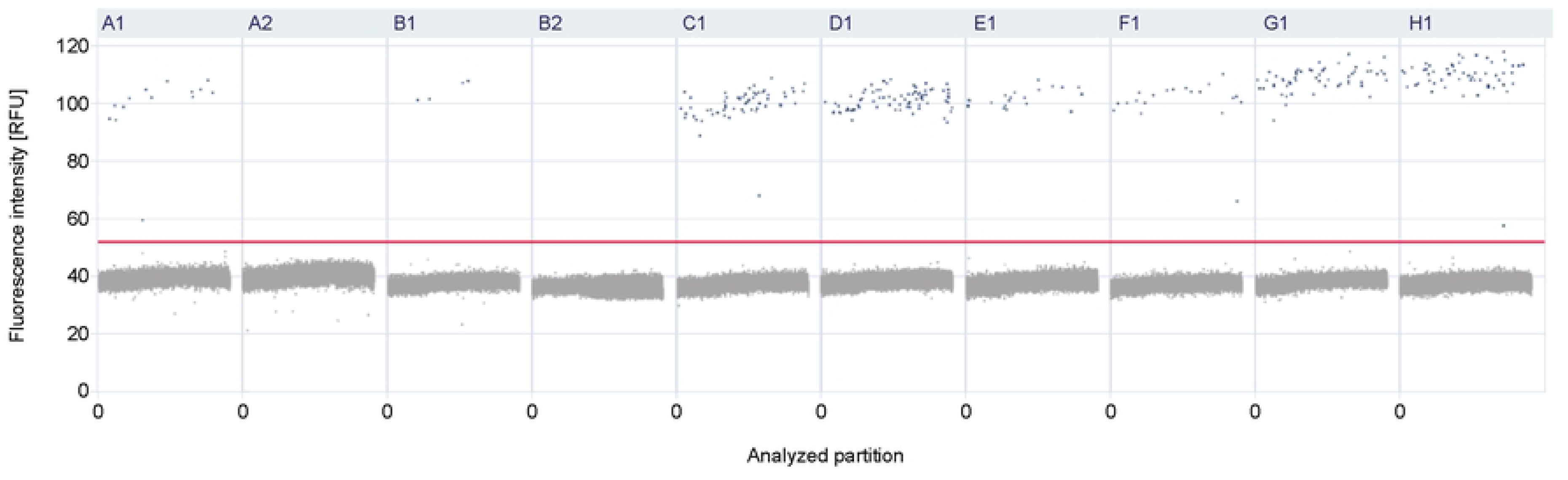
Optimization of HAV using dPCR. Examples of results from anti-HAV IgM-positive clinical samples that were used for optimization and verification. The red line represents the threshold, the gray area/dots under the threshold represent the negative partitions whereas blue dots above the threshold represent the positive partitions.

**Fig. 3.**
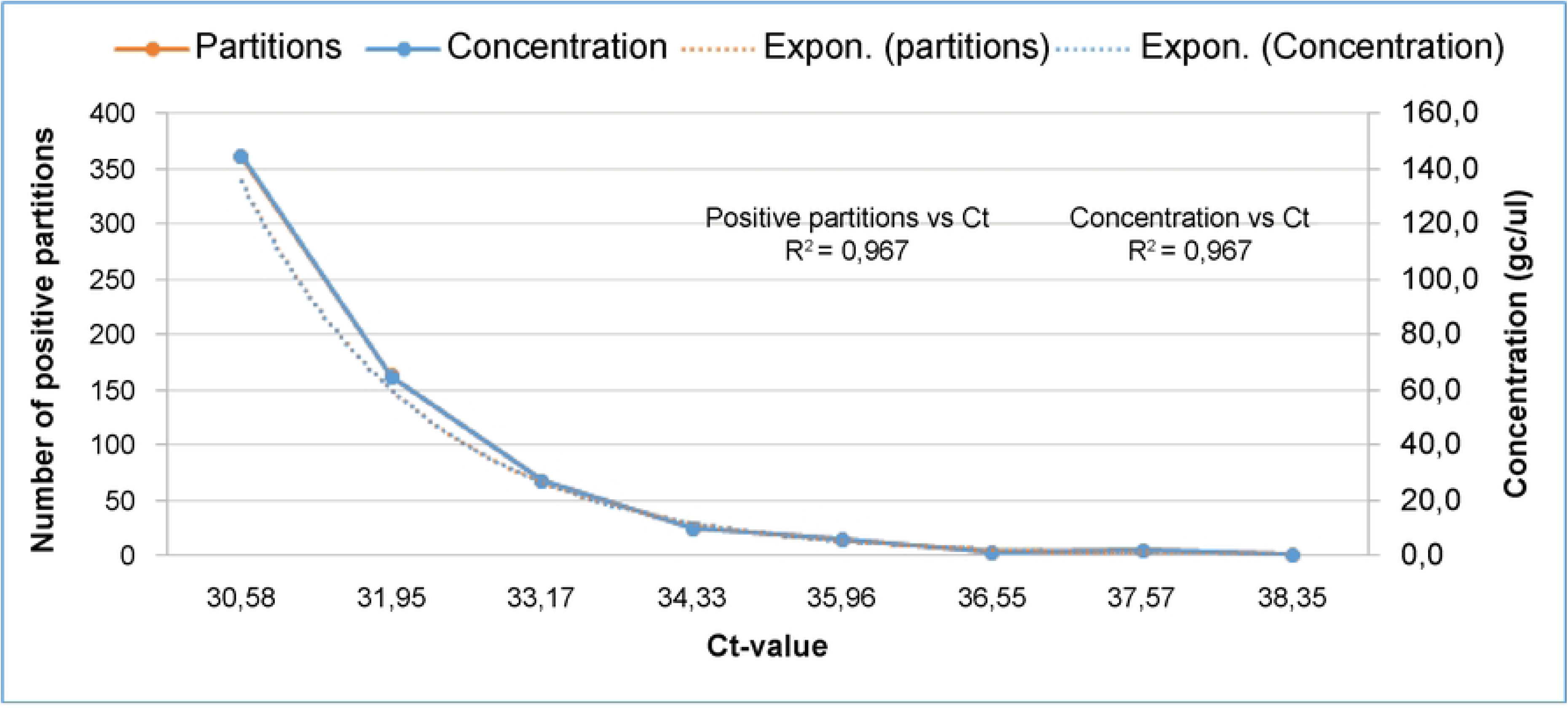
Limit of quantification comparing the number of partitions and concentration to ct-values. Two-fold dilutions were performed and results were plotted as an exponential curve. The orange points indicates the number of positive partitions and the blue points indicate the concentrations (genome copies) that correspond with the ct-values based on HAV detected for each dilution.

### 3.2. HAV detection in wastewater concentrates from 2021 to 2024

In total, 2013 wastewater samples were tested for HAV, of which 17.3% (349/2013) tested positive (Table 1). The majority of the positive samples had 1–5 partitions on dPCR (87.1%; 304/349) which was equivalent to 2.0–2.7 log gc/mL, followed by 6–10 partitions (5.7%; 20/349) which was equivalent to 2.8–3.0 log gc/mL (Fig. 4).

**Fig. 4.**
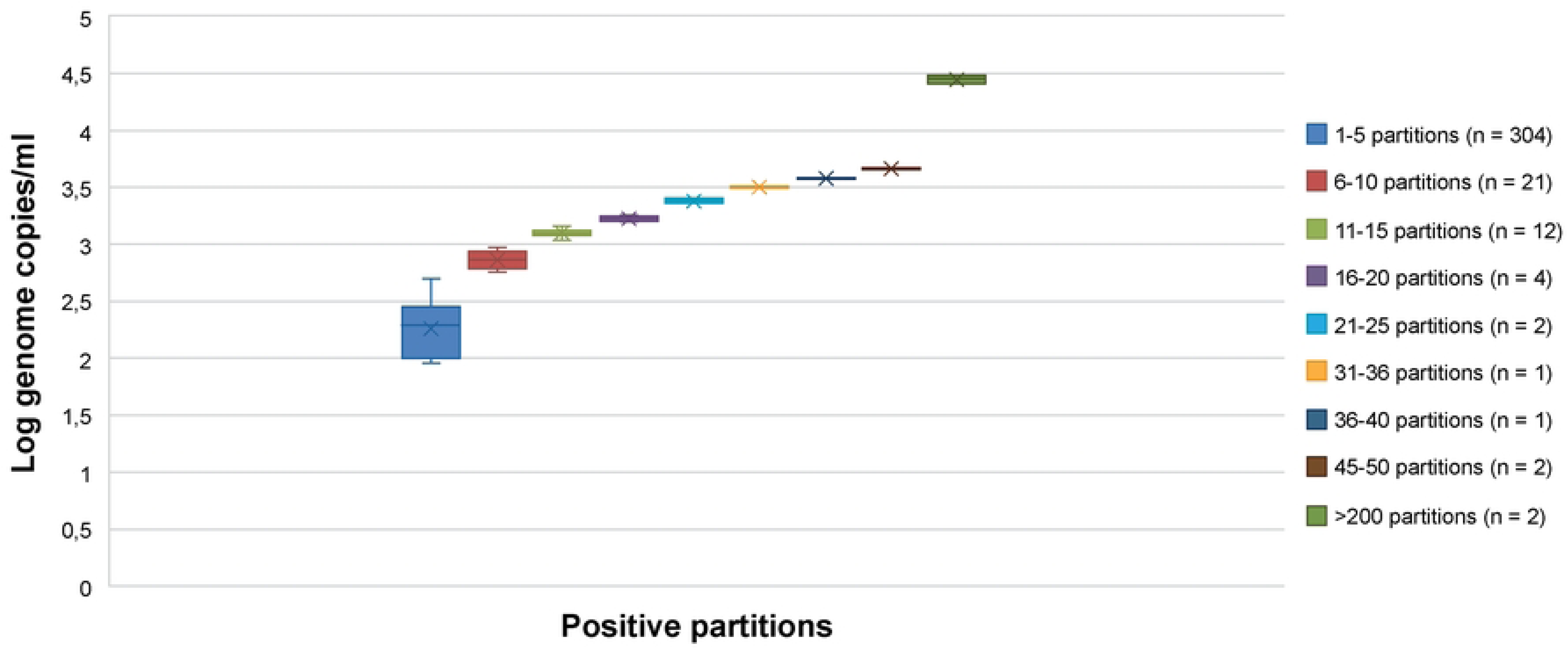
Genome copies/mL for samples with specified numbers of positive dPCR partitions, for all wastewater samples that tested positive for HAV. The legend displays the range of partitions for each of the box and whiskers plotted and in brackets is the number of samples that had a specific range of partitions.

**Table 1.** Results of wastewater samples tested for hepatitis A, from 2021 to 2023.

| **Results** | **Number of samples (N=2013)** | **Number of positive partitions on dPCR** | **Genome copies/ml** | **Equivalent log-transformed genome copies/μl** |
| --- | --- | --- | --- | --- |
| **Negative** | 1664 (82.7%) | 0 | 0 | 0 |
| **Positive** | 349 (17.3%) | 1-306 | 109-37410 | 2.0 - 4.5 |

HAV was detected in wastewater samples from several provinces of South Africa (Table 2). The majority of the samples tested were from Gauteng Province (70.1%; 1411/2013), with a positivity rate of 17.1%. When determining the positivity rates within each province, Limpopo (33.3%, 4/12) and the Western Cape (32.4%, 22/68) showed the highest intraprovince positivity.

**Table 2.** Wastewater samples tested for HAV across provinces in South Africa, from 2021 to 2024.

| **Provinces/Districts** | **Period** | **Total tested** | **Positive** | **Positivity rate (%)** |
| --- | --- | --- | --- | --- |
| **Eastern Cape** | Apr 2021-Mar 2024 | 173 | 26 | 15,0 |
| **Free State** | Aug 2021-Mar 2024 | 169 | 32 | 18,9 |
| **Gauteng** | Feb 2021-Mar 2024 | 1411 | 241 | 17,1 |
| **KwaZulu-Natal** | May 2021-Mar 2024 | 135 | 15 | 11,1 |
| **Limpopo** | Jan 2024-Mar 2024 | 12 | 4 | 33,3 |
| **Mpumalanga** | Jan 2024-Mar 2024 | 10 | 2 | 20,0 |
| **North West** | Oct 2023-Mar 2024 | 21 | 5 | 23,8 |
| **Northern Cape** | Aug 2021-May 2022 | 14 | 2 | 14,3 |
| **Western Cape** | Apr 2021-Mar 2024 | 68 | 22 | 32,4 |
| **All provinces** | Feb 2021-Mar 2024 | 2013 | 349 | 17,3 |

We further compared the HAV data from wastewater to clinical samples (Fig. 5). Overall, the proportion of HAV-positive wastewater samples was higher than that of anti-HAV IgM-positive samples. Western Cape province (5.1-9.0%) had the highest proportion of anti-HAV IgM positives. Although fewer wastewater sites were included in this province, the proportion of HAV-positive samples was >21%. In Gauteng, the proportion of anti-HAV IgM-positive cases was greatest in the City of Tshwane and Ekurhuleni districts (<1.0%), followed by the City of Johannesburg (1.1-2.0%). The proportion of HAV that tested positive in wastewater was the highest in the cities of Johannesburg and Ekurhuleni districts (>13%).

**Fig. 5.**
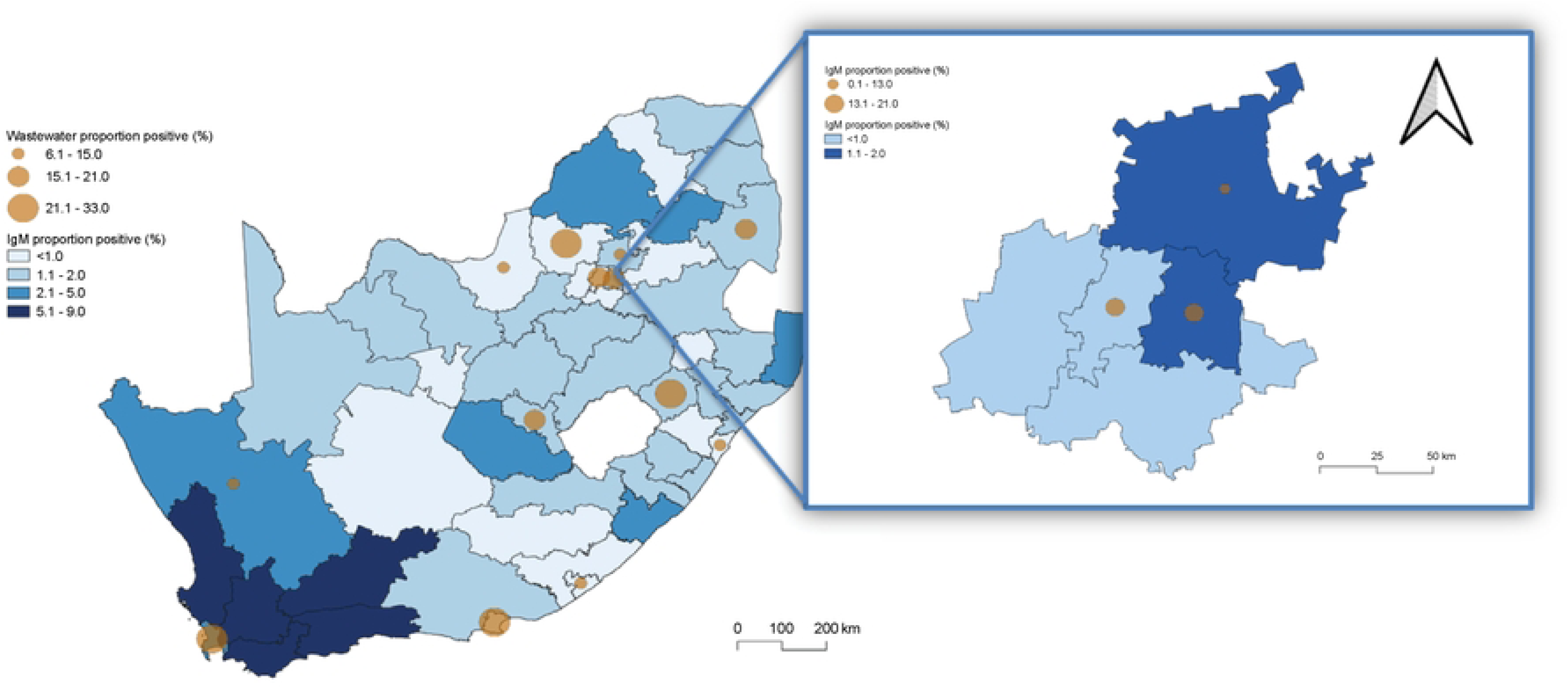
Proportion of HAV positive across South Africa, clinical vs wastewater. Brown circles represent the proportion of samples that tested positive for HAV, the larger the circle the higher the positivity. The shades of blue for each province (and districts within the province) represent the proportion of patients that were anti-HAV IgM positive: the darker the area is, the higher is the proportion of positivity. The figure on the right is a projection of the proportion positive from wastewater and clinical data in the Gauteng province.

### 3.3. Correspondence with clinical HAV surveillance data

Between epidemiological Week 7, 2021, and Week 9, 2024, 442 628 clinical samples were tested for anti-HAV IgM by NHLS laboratories. Two percent (9 098/ 442 628) of the samples were IgM-positive. During the same period, 17.3% (349/ 2013) of wastewater samples tested positive for HAV.

A total of 643 epidemiological week–district pairs were identified for clinical and wastewater samples in the districts where wastewater testing was conducted (Table 3). Among these, 26.1% (168/643) had HAV detected in at least one wastewater sample and one clinical sample (positive concordance). In 20.8% (134/643) of the epidemiological week–district pairs, no wastewater samples or clinical specimens tested positive for HAV (negative concordance). Wastewater week–district pairs were infrequently positive when clinical samples from the same week and district were negative (4.4%; 28/643; discordant wastewater positive). However, at least one clinical case of HAV was reported in 48.7% (313/643) of the districts during an epidemiological week when no wastewater samples tested positive for HAV (discordant wastewater negative; Table 3). When the definition of a positive week– district pair was expanded to include clinical cases reported in the epidemiological weeks before and after the wastewater sample testing, the number of concordant pairs increased (Table 4).

**Table 3.**
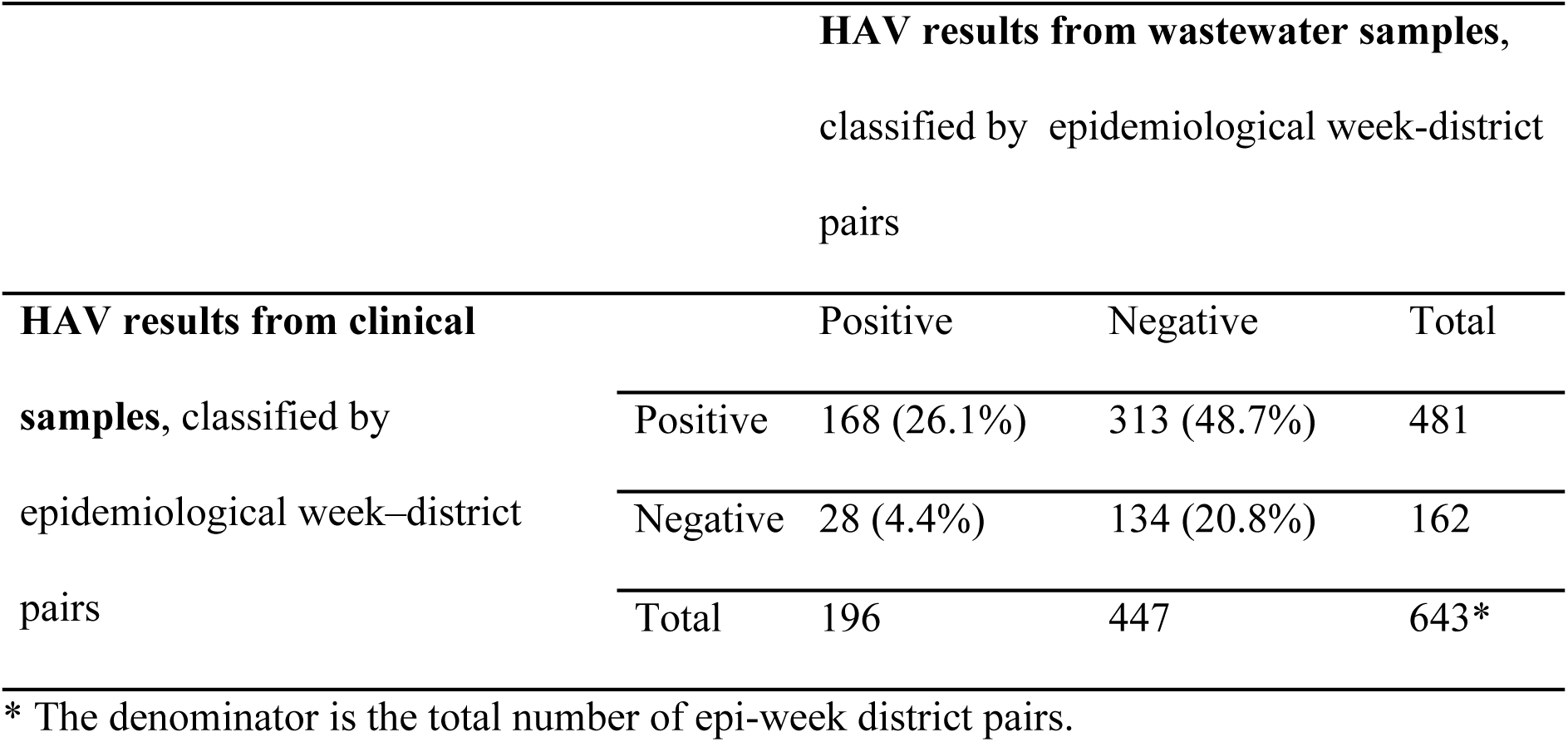
HAV results from clinical and wastewater samples grouped by corresponding epidemiological week: districts from epidemiological Week 7, 2021, to epidemiological Week 9, 2024.

**Table 4.** HAV results from clinical and wastewater samples grouped by corresponding epidemiological week: districts from epidemiological Week 7, 2021, to epidemiological Week 9, 2024.

| Same Epidemiological Week + Clinical Results 1 Week After and 2 Weeks After |  |  |  |  |
| --- | --- | --- | --- | --- |
|  |  | Wastewater Results |  |  |
|  |  | Positive | Negative | Total |
| Clinical Results (IgM) | Positive (IgM) | 191 (29.7%) | 402 (62.5%) | 593 |
|  | Negative (IgM) | 5 (0.8%) | 45 (7%) | 50 |
|  | Total | 196 | 447 | 643 |
| Same Epidemiological Week + Clinical Results 1 Week Before and 2 Weeks Before |  |  |  |  |
|  |  | Wastewater Results |  |  |
|  |  | Positive | Negative | Total |
| Clinical Results (IgM) | Positive (IgM) | 190 (29.5%) | 408 (63.5%) | 598 |
|  | Negative (IgM) | 6 (0.9%) | 39 (6.1%) | 45 |

|  | Total | 196 | 447 | 643 |
| --- | --- | --- | --- | --- |

With a positive week–district definition expanded to include clinical cases that were reported in epidemiological weeks before and after wastewater sample testing. *A positive epidemiological week–district pair is defined as the presence of at least one wastewater sample that tested positive in a given epidemiological week, or at least one clinical sample that tested positive for HAV 1 or 2 weeks before or after the wastewater sample tested positive, to accommodate for prolonged shedding (of up to 4 weeks) in HAV-infected individuals.

## 4. Discussion

HAV is shed for up to 1–3 weeks through the stool of infected individuals and is highly transmissible in the first 2 weeks before symptom onset, during which the highest viral concentration is attained in the stool [14]. HAV detection in wastewater samples can enhance our ability to monitor largely infected populations by collecting samples from a limited number of sites, including wastewater treatment plants, because waste from multiple households, clinics, and hospitals passes through a single site [18]. The incidence of HAV has been decreasing, and the average age of infected individuals is increasing globally, with most infections still prevalent in younger children [6]. Young children infected with the virus are often asymptomatic, do not seek healthcare, and are not reported [5]. We showed that a wastewater-based surveillance approach can enable the identification of HAV that goes undetected and thus provide an early-detection and warning system.

We adopted an ultrafiltration technique to concentrate and enrich the wastewater samples. Although relatively expensive, this approach requires no preconditioning steps and has been previously used to detect HAV in Africa [32]. A systematic review of the global prevalence of HAV in wastewater from 1986 to 2020 reported that the overall HAV prevalence was 31.4% in untreated wastewater, with viral concentrations of up to 3.7 × 10^10^ gc/L, which was higher than concentrations detected in other water sources [33]. In South Africa, HAV was detected in 43.1% of samples collected from various sites along the Buffalo River [21].

Another study described high levels of HAV detected at concentrations of 1.34 × 10^5^ to 3.7 × 10^10^ gc/L in sewage with an 80% detection rate in five wastewater treatment plants in South Africa during 2015 and 2016 [34]. The prevalence of HAV in South African-treated sewage and surface water samples ranges from 4% to 37% and 16% to 76%, with an estimated average detection rate of 15% (95% CI 1–29) and 51% (95% CI 21–80), respectively [10].

Detecting HAV in water matrices involves various detection methods, including the most frequently used conventional RT-PCR (54.5%), followed by real-time PCR (37.5%) [33]. dPCR offers greater precision in comparison to qPCR and is far simpler to use for copy-number quantification owing to its binary nature wherein the partitions are counted as positive or negative [35]. To detect and quantify HAV from wastewater samples, we successfully optimized a dPCR method that was developed using previously published primers to detect and quantify HAV from wastewater samples. Similarly, although our wastewater samples had lower HAV concentrations, HAV was detected in the untreated water sources. This could be attributed to the time of viral shedding or dilution of the viral concentration diluted during flow from the distributaries to the wastewater site. Our results showed that the detected viral concentrations of HAV are a good representation of the epidemiological distribution of HAV infection in the surrounding population, where the virus is excreted from infected individuals .

Takuissu et al. [2023], in a systematic review, showed that low-income economies had the highest HAV prevalence (29.0%), and Africa had the highest HAV prevalence. Varying proportions of HAV have been detected in wastewater samples collected from different provinces of South Africa. In our study, although the number of wastewater samples collected varied across the different provinces in South Africa, in the samples collected from the Eastern Cape, Free-State, Gauteng, Kwa-Zulu Natal and Western Cape provinces the proportion of HAV in wastewater samples was generally higher compared to the proportion of anti-HAV IgM reported from clinical data. The primary wastewater sites were located in Gauteng Province, in the city of Tshwane, Johannesburg, and Ekurhuleni districts. In these districts, the proportion of HAV in wastewater samples was >10 times greater than the proportion of anti-HAV IgM from clinical samples. Our findings suggested that a large proportion of HAV clinical cases were undetected, which could be attributable to asymptomatic HAV-infected patients who would not have sought healthcare services [18].

Further comparison of the clinical data to wastewater data by epidemiological week and district showed some concordance (26.1%) between the positive HAV results. However, <50% of the samples showed discordant results on comparing HAV-positive results from clinical samples to HAV-negative results from wastewater samples. This finding could be a result of the larger number of clinics and hospitals tested across South Africa during the study period compared with the number of sites tested for wastewater.

### 4.1. Limitations

First, a limitation of this study was the low number of samples collected from wastewater sites in provinces other than Gauteng Province, which made it difficult to determine the true prevalence of HAV in South Africa from wastewater samples. Second, the number of sites with clinical cases was much more extensive than that of wastewater sites. Our study demonstrated the feasibility of using wastewater to complement clinical surveillance in South Africa. Further comparisons with clinical data are required to establish the significance of HAV detection in wastewater. Third, the enrichment method may have contributed to lower viral concentrations, and alternative methods should be tested to improve viral concentration.

Lastly, the concentration of HAV detected in the wastewater samples may be lower than expected because the samples were tested retrospectively, and freeze/thawing may have resulted in RNA degradation and reduced viral loads. Prospective testing of the samples will eliminate this factor and enable the detection of accurate virus concentrations.

## 5. Conclusion

Low- to middle-income countries with restricted access to appropriate resources can benefit from wastewater surveillance for monitoring HAV incidence in asymptomatic and symptomatic populations and identifying outbreaks and potential sources of transmission. To expand on our current data, future work is required to compare wastewater data with clinical surveillance data to facilitate the appropriate interpretation of the results; next-generation sequencing should be performed to determine the epidemiological significance of HAV in South Africa.

## Data Availability

All relevant data are within the manuscript and its Supporting Information files

## Acknowledgments

Editorial support, in the form of medical writing, assembling tables and creating high-resolution images based on authors’ detailed directions, collating author comments, copyediting, fact checking, and referencing, was provided by Editage, Cactus Communications.

## Notes

### Competing Interest Statement

The authors have declared no competing interest.

### Funding Statement

The funder: Bill and Melinda Gates Foundation (https://www.gatesfoundation.org/), grant (INV-049271), and (INV-050051); MY and KM were the PIs. The funders had no role in the data collection and analysis, decision to publish, or preparation of the manuscript

## References

1. World Health Organization. https://www.who.int/health-topics/hepatitis#tab=tab_2 (accessed 29 May 2023). 2023. Hepatitis.

2. Nainan OV, Xia G, Vaughan G, Margolis HS. Diagnosis of hepatitis a virus infection: a molecular approach. Clin Microbiol Rev. 2006;19: 63–79. doi:10.1128/CMR.19.1.63-79.2006.

3. Shin EC, Jeong SH. Natural history, clinical manifestations, and pathogenesis of hepatitis A. Cold Spring Harb Perspect Med. 2018;8: a031708. doi:10.1101/cshperspect.a031708.

4. Cao G, Jing W, Liu J, Liu M. The global trends and regional differences in incidence and mortality of hepatitis A from 1990 to 2019 and implications for its prevention. Hepatol Int. 2021;15: 1068–1082. doi:10.1007/s12072-021-10232-4.

5. Abutaleb A, Kottilil S. Hepatitis A: Epidemiology, natural history, unusual clinical manifestations, and prevention. Gastroenterol Clin North Am. 2020;49: 191–199. doi:10.1016/j.gtc.2020.01.002.

6. Jacobsen KH. Globalization and the changing epidemiology of hepatitis A virus. Cold Spring Harb Perspect Med. 2018;8: a031716. doi:10.1101/cshperspect.a031716.

7. NICD. Communicable Diseases Communique. January 2023, vol.22(1). Vol. 22. 2023.

8. South African Department of Health. 2019. Hepatitis A. National guidelines for the management of viral hepatitis. https://knowledgehub.health.gov.za/system/files/elibdownloads/2023-04/SA%20NDOH_Viral%20Hepatitis%20guidelines%20final.pdf. (Accessed{24/09/2024}).

9. Prabdial-Sing N, Motaze V, Manamela J, McCarthy K, Suchard M. Establishment of Outbreak Thresholds for Hepatitis A in South Africa Using Laboratory Surveillance, 2017-2020. Viruses. 2021;13: 2470. doi:10.3390/v13122470.

10. Kuodi P, Patterson J, Silal S, Hussey GD, Kagina BM. Characterisation of the environmental presence of hepatitis A virus in low-income and middle-income countries: a systematic review and meta-analysis. BMJ Open. 2020;10: e036407. doi:10.1136/bmjopen-2019-036407.

11. Enoch A, Hardie DR, Hussey GD, Kagina BM. Hepatitis A seroprevalence in Western Cape Province, South Africa: Are we in epidemiological transition. S Afr Med J. 2019;109: 314–318. doi:10.7196/SAMJ.2019.v109i5.13410.

12. du Plessis NM, Haeri Mazanderani A, Motaze NV, Ngobese M, Avenant T. Hepatitis A virus seroprevalence among children and adolescents in a high-burden HIV setting in urban South Africa. Sci Rep. 2022;12: 20688. doi:10.1038/s41598-022-25064-x.

13. Haeri Mazanderani A, Motaze NV, McCarthy K, Suchard M, du Plessis NM. Hepatitis A virus seroprevalence in South Africa - Estimates using routine laboratory data, 2005- 2015. PLoS One. 2019;14: e0216033. doi:10.1371/journal.pone.0216033.

14. EMRO WHO. WHO. Polio Eradication Initiative, Surveillance.

15. World Health Organization [Internet]. 2022. WHO. Available from: https://www.who.int/health-topics/coronavirus

16. Rachida S, Matsapola PN, Wolfaardt M, Taylor MB. Genetic characterization of a novel hepatitis a virus strain in irrigation water in South Africa. J Med Virol. 2016;88: 734–737. doi:10.1002/jmv.24370.

17. Street R, Malema S, Mahlangeni N, Mathee A. Wastewater surveillance for Covid-19: An African perspective. Science of the Total Environment. 2020;743: 1–3. doi: 10.1016/j.scitotenv.2020.140719

18. Hellmér M, Paxéus N, Magnius L, Enache L, Arnholm B, Johansson A, et al. Detection of pathogenic viruses in sewage provided early warnings of hepatitis A virus and norovirus outbreaks. Appl Environ Microbiol. 2014;80: 6771–6781. doi:10.1128/AEM.01981-14.

19. Sattar SA, Jason T, Bidawid S, Farber J. Foodborne spread of hepatitis A: Recent studies on virus survival, transfer and inactivation. Can J Infect Dis. 2000;11: 159–163. doi:10.1155/2000/805156.

20. Vaughan G, Goncalves Rossi LM, Forbi JC, de Paula VS, Purdy MA, Xia G, et al. Hepatitis A virus: host interactions, molecular epidemiology and evolution. Infect Genet Evol. 2014;21: 227–243. doi:10.1016/j.meegid.2013.10.023.

21. Chigor VN, Okoh AI. Quantitative RT-PCR detection of hepatitis A virus, rotaviruses and enteroviruses in the Buffalo River and source water dams in the Eastern Cape Province of South Africa. Int J Environ Res Public Health. 2012;9: 4017–4032. doi:10.3390/ijerph9114017.

22. Adefisoye MA, Nwodo UU, Green E, Okoh AI. Quantitative PCR detection and characterisation of human adenovirus, rotavirus and hepatitis A virus in discharged effluents of two wastewater treatment facilities in the Eastern Cape, South Africa. Food Environ Virol. 2016;8: 262–274. doi:10.1007/s12560-016-9246-4.

23. World Health Organization. World Health Statistics 2020. 2022. 1–77 p.

24. Osuolale O, Okoh A. Incidence of human adenoviruses and Hepatitis A virus in the final effluent of selected wastewater treatment plants in Eastern Cape Province, South Africa. Virol J. 2015;12: 98. doi:10.1186/s12985-015-0327-z.

25. CDC. National Wastewater Surveillance System (NWSS). 2020.

26. Silveira Carneiro J, Equestre M, Pagnotti P, Gradi A, Sonenberg N, Perez Bercoff R. 5’ UTR of hepatitis A virus RNA: mutations in the 5’-most pyrimidine-rich tract reduce its ability to direct internal initiation of translation. J Gen Virol. 1995;76 (Pt 5): 1189–1196. doi:10.1099/0022-1317-76-5-1189.

27. Liu GD, Hu NZ, Hu YZ. Full-length genome of wild-type hepatitis A virus (DL3) isolated in China. World J Gastroenterol. 2003;9: 499–504. doi:10.3748/wjg.v9.i3.499.

28. Wu S, Nakamoto S, Kanda T, Jiang X, Nakamura M, Miyamura T, et al. Ultra-deep sequencing analysis of the hepatitis A virus 5’-untranslated region among cases of the same outbreak from a single source. Int J Med Sci. 2014;11: 60–64. doi:10.7150/ijms.7728.

29. Prajapati B, Rathore D, Joshi C, Joshi M. Digital PCR: A partitioning-based application for detection and surveillance of SARS-CoV-2 from sewage samples. Methods Mol Biol. 2023;2967: 1–16. doi:10.1007/978-1-0716-3358-8_1.

30. Costafreda MI, Bosch A, Pintó RM. Development, evaluation, and standardization of a real-time TaqMan reverse transcription-PCR assay for quantification of hepatitis A virus in clinical and shellfish samples. Appl Environ Microbiol. 2006;72: 3846–3855. doi:10.1128/AEM.02660-05.

31. Iwu-Jaja C, Ndlovu NL, Rachida S, Yousif M, Taukobong S, Macheke M, et al. The role of wastewater-based epidemiology for SARS-CoV-2 in developing countries: Cumulative evidence from South Africa supports sentinel site surveillance to guide public health decision-making. Sci Total Environ. 2023;903: 165817. doi:10.1016/j.scitotenv.2023.165817.

32. Osunmakinde CO, Selvarajan R, Sibanda T, Mamba BB, Msagati TAM. Overview of trends in the application of metagenomic techniques in the analysis of human enteric viral diversity in Africa’s environmental regimes. Viruses. 2018;10: 429. doi:10.3390/v10080429.

33. Takuissu GR, Kenmoe S, Ebogo-Belobo JT, Kengne-Ndé C, Mbaga DS, Bowo-Ngandji A, et al. Occurrence of hepatitis A virus in water matrices: a systematic review and meta-analysis. Int J Environ Res Public Health. 2023;20: 1054. doi:10.3390/ijerph20021054.

34. Rachida S, Taylor MB. Potentially infectious novel hepatitis A virus strains detected in selected treated wastewater discharge sources, South Africa. Viruses. 2020;12: 1468. doi:10.3390/v12121468.

35. Whale AS, Huggett JF, Cowen S, Speirs V, Shaw J, Ellison S, et al. Comparison of microfluidic digital PCR and conventional quantitative PCR for measuring copy number variation. Nucleic Acids Res. 2012;40: e82. doi:10.1093/nar/gks203.

